# Comparative Study of Serum Calcium, 24-hour Urinary Calcium, and Body Mass Index in Patients with Urolithiasis and Their First-Degree Relatives

**DOI:** 10.1101/2025.09.30.25337019

**Authors:** Kidirali Karimbayev, Zhanar Ashirbayeva, Zhanat Nakipova, Yerbol Tulezhanov, Mirzakarim Alchinbayev, Daniyar Ustimirov, Narymzhan Nakisbekov

## Abstract

**Background:** Urolithiasis is a major public health concern worldwide, with a rising incidence in Kazakhstan. Early detection of metabolic abnormalities among first-degree relatives of affected patients may improve prevention strategies.

**Objectives:** To evaluate the diagnostic significance of hypercalcemia, hypercalciuria, and body mass index (BMI) in patients with urolithiasis and their first-degree relatives as potential predictors of stone formation.

**Methods:** A retrospective study was conducted including 180 patients with radiologically confirmed urolithiasis and 315 of their siblings. Serum calcium and 24-hour urinary calcium were measured using standard biochemical analyzers. Hypercalcemia was defined as serum calcium >2.65 mmol/L, and hypercalciuria as urinary calcium excretion >7.5 mmol/24 h in men and >6.5 mmol/24 h in women. BMI was categorized according to WHO criteria. Statistical analyses included descriptive statistics and chi-square testing (p<0.05). Written informed consent was obtained from all participants; for minors, consent was provided by their parents or legal guardians.

**Results:** Hypercalcemia was observed in 82 patients (45.6%) and hypercalciuria in 78 (43.3%). Among siblings, hypercalcemia occurred in 32.7% and hypercalciuria in 17.1%, indicating subclinical metabolic alterations. Simultaneous hypercalcemia and hypercalciuria were detected in 64.5% of siblings of patients with hypercalcemia. Obesity/overweight was found in 80% of patients but only in 12% of siblings. Male patients with hypercalcemia showed particularly high concordance with their brothers’ calcium abnormalities.

**Conclusions:** Hypercalcemia, hypercalciuria, and elevated BMI are frequent abnormalities among patients with urolithiasis. A substantial proportion of siblings also exhibit latent metabolic risk factors, underscoring the role of genetic predisposition. Routine biochemical screening of first-degree relatives could aid in early detection and prevention of urolithiasis.

## Introduction

Urolithiasis remains one of the most frequent urological diseases worldwide, affecting 1–20% of the population depending on geography and ethnicity [1]. According to the Global Burden of Disease Study, in 2019 more than 115 million new cases were recorded globally [2]. In Kazakhstan, the incidence of urolithiasis increased from 75.1 per 100,000 in 2014 to 83.1 per 100,000 in 2023 [3]. Stone formation is a multifactorial process involving supersaturation of urine with salts, nucleation, growth, and aggregation of crystals [4,5]. Environmental factors, dietary habits, genetic predisposition, and metabolic abnormalities all contribute [6,7]. Among metabolic factors, hypercalciuria and hypercalcemia play central roles [8–10]. Hypercalciuria is found in 40–50% of recurrent stone formers [11], while hypercalcemia may indicate primary hyperparathyroidism or other endocrine disorders [12–15]. Not all patients with these disorders develop stones, suggesting multifactorial pathogenesis [16]. Obesity and overweight also increase lithogenic risk [6]. Randall’s plaques—calcium phosphate deposits in renal papillae—are important initiating sites for stone formation [17]. Given the high recurrence rate of urolithiasis (up to 50% within five years) [18–20], early identification of risk factors is essential [21–24].

## Materials and Methods

This retrospective study included 180 patients with radiologically confirmed urolithiasis treated between January 1, 2023, and July 30, 2025, as well as 315 of their siblings. During this period, the research team had full access to the medical records and related materials. The authors had official permission to access identifiable participant information as required for the study. Inclusion criteria: confirmed diagnosis, availability of biochemical tests, and consent to include relatives. Exclusion criteria: severe chronic kidney disease, endocrine disorders unrelated to calcium metabolism, refusal to participate. All subjects underwent anthropometric measurements, BMI was calculated and categorized according to WHO. Serum and 24-hour urinary calcium were measured. Hypercalcemia was defined as serum calcium >2.65 mmol/L, and hypercalciuria as urinary calcium excretion >7.5 mmol/24 h in men and >6.5 mmol/24 h in women. Data were grouped by sex and calcium status. Statistical analysis used descriptive statistics, chi-square testing, and 95% confidence intervals, with significance set at p<0.05. Written informed consent was obtained from all participants after they were provided with detailed information about the study and had the opportunity to review it. For underage participants, consent was obtained from their parents or legal guardians

## Results

Among the 180 patients, 110 (61.1%) were men and 70 (38.9%) women, mean age 44.8 years. In 165 (91.7%) urolithiasis was newly diagnosed, while 15 (8.3%) had recurrences. Stone localization: solitary renal stones in 29, staghorn/multiple in 19, and ureteral stones in 132. Hypercalcemia was found in 82 patients (45.6%), normocalcemia in 98 (54.4%), hypocalcemia in 17 (9.4%). Hypercalciuria was detected in 78 (43.3%). Overweight/obesity in 80% of patients. Among siblings (n=315), hypercalcemia in 103 (32.7%), hypercalciuria in 54 (17.1%). Simultaneous hypercalcemia and hypercalciuria in 64.5% of siblings of hypercalcemic patients. Most siblings (88%) had normal BMI; only 12% overweight/obese. Male patients with hypercalcemia showed particularly high concordance with their brothers’ calcium abnormalities. Overall, calcium abnormalities were significantly more frequent among patients than siblings (p<0.05).

## Discussion

This study confirms the high prevalence of hypercalcemia, hypercalciuria, and elevated BMI among patients with urolithiasis [6,8,11,13]. Importantly, a substantial proportion of siblings also displayed subclinical abnormalities, highlighting the role of genetic predisposition [17,18]. Hypercalcemia was present in 45.6% of patients, consistent with previous findings linking it to primary hyperparathyroidism and altered vitamin D metabolism [12–15]. Hypercalciuria was observed in 43.3% of patients, in agreement with international data [10,11]. Obesity was also strongly associated with calcium disturbances [6,16]. Our results demonstrate that urolithiasis is not solely an acquired condition, but reflects genetic, metabolic, and lifestyle interactions [19– 24]. Routine biochemical screening of first-degree relatives could identify at-risk individuals for early preventive interventions.

## Conclusions

Hypercalcemia, hypercalciuria, and elevated BMI are common abnormalities among patients with urolithiasis. A significant proportion of siblings also demonstrate subclinical metabolic risk factors. Routine biochemical screening of siblings, along with BMI assessment and lifestyle modifications, may help prevent urolithiasis.

## Data Availability

The data does not belong to a third party and the authors have permission to distribute the data.

## Ethics Committee

The study was conducted in accordance with the Declaration of Helsinki, and approved by the Ethics Committee Khoja Akhmet Yassawi International Kazakh-Turkish University (protocol code No. 9, 14 November 2022).

## Acknowledgments

The authors express their gratitude to the staff of the Clinical and Diagnostic Center of Khoja Akhmet Yassawi International Kazakh-Turkish University for their assistance in conducting this research.

## Author Contributions

Conceptualization: Alchinbayev MK, Karimbayev KK, Nakipova ZZ. Data collection: Karimbayev KK, Ashirbayeva ZM, Tulezhanov EN. Data analysis: Ustimirov DN, Nakisbekov NO, Karimbayev KK. Draft preparation: Karimbayev KK, Ashirbayeva ZM, Nakipova ZZ. Review and editing: Karimbayev KK, Nakipova ZZ. All authors approved the final version.

